# Mission imputable: Effects of missing data processing on infectious disease detection and prognosis

**DOI:** 10.1101/2025.02.15.25322351

**Authors:** Suravi Saha Roy, Ngoc Thi Nguyen, Agustin Zuniga, Fatemeh Sarhaddi, Eemil Lagerspetz, Huber Flores, Petteri Nurmi

## Abstract

**Background:** Missing data in medical datasets poses significant challenges for developing effective AI/ML pipelines. Inaccurate imputation can lead to biased results, reduced model performance, and compromised clinical insights. Understanding how different imputation methods affect AI/ML model performance is crucial for ensuring accurate clinical findings.

**Objective:** This study systematically investigates the effects of different imputation methods on AI/ML model performance and the clinical implications of these methods.

**Methods:** We investigate the impact of four different missing data strategies on the performance of common classification algorithms for analyzing medical data. The performance was evaluated based on sensitivity and specificity metrics for the tasks of predicting COVID-19 diagnosis and patient deterioration. We also perform feature analysis to understand the clinical implications the choice of imputation method has.

**Results:** Our findings show that the choice of imputation method significantly affects the performance of AI/ML techniques and the clinical conclusions drawn from the data. The optimal handling of missing values depends on (i) the composition of the features with missing values, (ii) the rate of missing values, and (iii) the pattern of the missing features. Using COVID-19 diagnosis and patient deterioration as representative examples of clinical tasks, our results indicate that MICE imputation yields the best overall performance, resulting in a 26% improvement in accuracy compared to baseline methods. Specifically, for predicting COVID-19 diagnosis, we achieved a sensitivity of 81% and specificity of 98%, while for patient deterioration, the sensitivity was 65% and specificity was 99%.

**Conclusion:** This study demonstrates the critical impact of missing data imputation on AI/ML model performance and the clinical insights derived from these models. Our findings underscore the importance of selecting appropriate imputation techniques tailored to the specific characteristics of medical data to ensure accurate and reliable AI/ML predictions.

## Introduction

Artificial Intelligence (AI) and machine learning (ML) methods have significant potential to alleviate pressure on healthcare systems by offering cost-effective and efficient solutions for supporting the analysis, detection and care of health conditions, thereby reducing reliance on time-consuming and complex clinical procedures. Indeed, ML techniques have demonstrated considerable promise in enhancing the diagnosis of various conditions, including cancer [1], cardiovascular diseases [2], lung infections [3], and diabetes onset [4], among others. However, the development of effective machine learning pipelines is often challenged by the heterogeneous nature of medical data, which is frequently characterized by high sparsity and substantial amounts of missing values [5]. If these issues are not adequately addressed, they can significantly impact the accuracy of machine learning models and the clinical insights derived from the data. For instance, Cismondi et al. [6] demonstrated an 11% difference in accuracy for detecting septic shocks based on whether missing data was appropriately handled or omitted.

Typically, methods for handling missing data are tailored to individual studies, resulting in limited insights regarding best practices and procedures. Recent research has explored various imputation methods [7] for both general datasets and health-specific datasets [8]. Other studies have reviewed different imputation techniques applied to healthcare applications based on clinical datasets for various diseases [9, 10], comparing the effectiveness of these methods. While these works provide valuable overviews of imputation methods across several healthcare domains, comprehensive and systematic evaluations of these techniques, particularly for infectious diseases, remain lacking. A detailed understanding of the advantages and disadvantages of different procedures for handling missing data, as well as their impact on the performance of AI and ML methods, is essential for developing reusable and effective pipelines that produce medically accurate clinical insights.

The COVID-19 pandemic underscored the importance of robust AI/ML applications to support healthcare and reduce the burden of health professionals. The use of AI/ML to detect COVID-19 infections, or predicting potential patient deterioration, are two representative examples of tasks where AI/ML methods operating on medical data can be highly useful. Indeed, since the onset of the COVID-19 pandemic, there has been a significant interest in developing machine learning methods to predict COVID-19 risks, infections, and outcomes [11]. These efforts range from analyses using chest x-ray images [12] to studies that take advantage of laboratory tests [13, 14] or electronic health records [15]. While these have shown promising results, the studies have been highly customized without offering insights into the effects of imputation techniques on machine learning pipelines or the clinical insights drawn from these analyses. Indeed, practically all existing studies are affected by significant amounts of missing data and high data sparsity, yet the methods for handling missing data are largely determined on a case-by-case basis, often without a comprehensive understanding of their clinical effects or overall impact.

This paper contributes a comprehensive and systematic analysis of the effects of different missing data strategies on AI/ML performance, with a particular focus on the clinical implications of these methods. We investigate the effects of four different strategies (no imputation and three different imputation methods) within a cost-effective pipeline for COVID-19 diagnosis and prognosis (i.e., a methodological process for early detection of COVID-19 infections and patient deterioration). Our analysis utilizes the openly available Hospital Israelita Albert Einstein dataset [16], which contains medical records from 1591 patients. Each record contains values for up to 106 different laboratory tests, including COVID-19 status and information about possible patient admissions to semi-intensive or intensive care units.

Our results demonstrate that the performance of machine learning models is heavily influenced by how missing data is handled, with improvements of up to 26 percentage points achievable through the incorporation of appropriate imputation techniques. The optimal approach to handling missing data depends on the characteristics of the missing data itself, including feature type, frequency, and pattern. Generally, ignoring missing data (i.e., no imputation) results in the poorest performance, while multiple imputation (multivariate imputations by chained equations, MICE) offers the best outcomes in most scenarios. We also analyze the importance of different features and highlight how the most significant predictors vary depending on the data processing method. For instance, when no imputation is performed, patient age quantile emerges as the best predictor for patient deterioration, whereas different viral tests and blood test features become more critical when the data is imputed. This underscores that imputation not only affects the performance of machine learning algorithms but can also lead to different clinical insights. Through our analysis, we also contribute an improved AI/ML pipeline for early screening, detecting COVID-19 positive patients with 81% sensitivity and 98% specificity, and in triaging by predicting patient deterioration with 65% sensitivity and 99% specificity.

Our work offers novel insights into the effects of imputation techniques, demonstrating the importance of missing data processing in the design of effective AI/ML pipelines. Researchers and data scientists can leverage our findings to improve the design of machine learning pipelines, while clinicians and medical professionals can gain more robust insights into the factors affecting AI performance. Additionally, we extend previous works by offering an effective, low-cost method for early screening and triaging based on laboratory tests. Clinicians can employ our methods for early screening of suspected COVID-19 patients, triaging the necessary medical procedures, and as a secondary diagnostic tool for prioritizing patient treatments using low-cost, readily available laboratory test results.

## Methods

### Dataset

In this study, we used an open-source dataset made publicly available by the Hospital Israelita Albert Einstein in São Paulo, Brazil through Kaggle [16]. The dataset was made anonymized by the publisher, following international guidelines, which ensures that no personally identifiable data is available. Therefore, no additional ethical approval was required to use it. We accessed and downloaded the dataset in April 2022 to develop our method for early COVID-19 screening and triaging, and to conduct our analysis on the effects of imputation methods. In total, 1591 patient records were used in this study, which consist of anonymized data from patients tested for COVID-19 using RT-PCR (positive or negative), admission decisions for positive cases (regular ward, semi-/intensive care unit), and 106 laboratory tests collected during their visit to the emergency room. The data has significant skew in outputs (approximately 9% of the samples are positive for COVID-19 tests) and a large number of missing laboratory test values, with 77.25% of the samples missing at least one laboratory test result.

The use of this real-world medical dataset ensures that our data is representative of regular clinical practices and incorporates clinical data from laboratory tests on suspected COVID-19 patients. This allows us to capture the relationships between the laboratory test outcomes, COVID-19 diagnosis, and patient deterioration. Note that the full release of the dataset contains 5644 records, but 71.81% of these (4053 samples) lack values for 20 laboratory tests considered in our analysis and hence, these records were omitted.

### Data analysis

The AI/ML pipeline used in our method for early COVID-19 screening and triaging and investigating the effects of different imputation methods on machine learning models is shown in Figure 1. In the pre-processing step, input data that consist of COVID-19 test, laboratory tests, and patient deterioration data are cleaned to remove empty records and to transform the features into numerical values. Then in the feature pruning step, the dimensionality of the data is reduced by selecting the representative features and removing other dependent features. The imputation step is for imputing the data using one of the imputation techniques considered in the study (no imputation, single imputation, multiple imputation KNN, and multiple imputation MICE). Finally, in the modeling step, different classifiers are trained and evaluated to predict COVID-19 diagnosis and patient deterioration. The classifiers include logistic regression (LR), random forest, support vector machine (SVM), and k-nearest neighbors (KNN). These steps are described in detail in the following sections.

**Fig 1.**
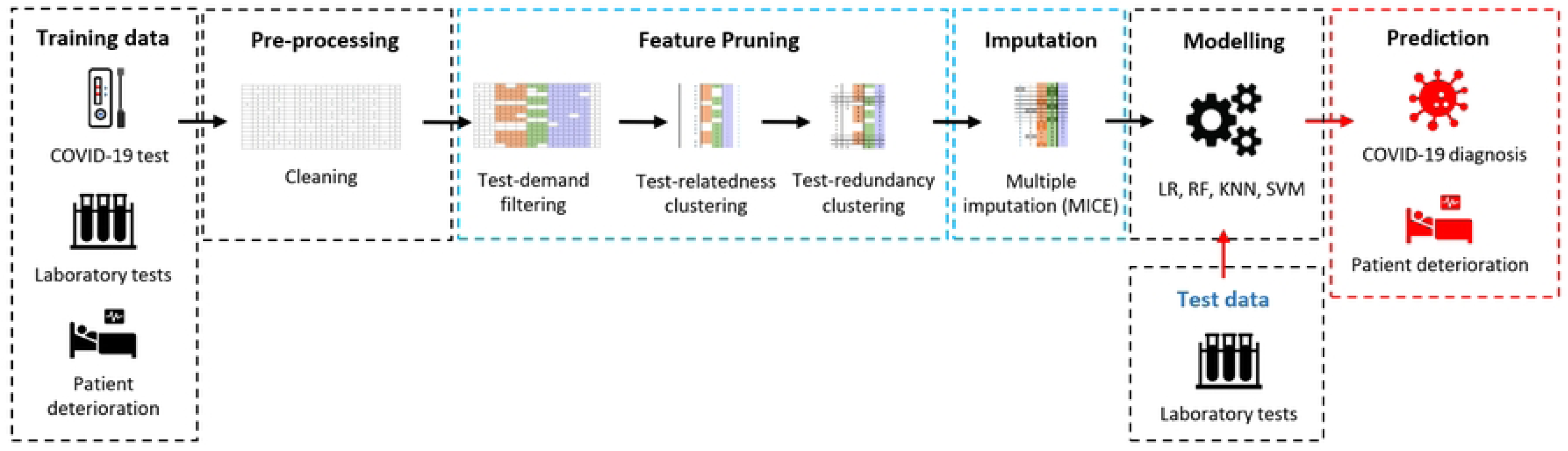
Missing data imputation pipeline through feature pruning and missing value imputation.

### Feature analysis

Medical practices and procedures govern which medical tests and procedures a specific patient undergoes. As these vary across patients, there are certain commonalities but also variations across patients. Accounting for these variations in procedures can be used to alleviate data sparsity. Specifically, the availability of a particular test variable depends on three interlinked factors: (i) *demand* for specific test procedures, i.e., whether a patient is required to undergo a specific laboratory test; (ii) *relatedness*, i.e., several tests monitor factors that are closely correlated with another test and it suffices to use of the tests; and (iii) *redundancy*, i.e., tests may measure the same aspect and be candidates for replacement. We take advantage of these three criteria to reduce feature space and to reduce biases and inaccuracies in the analysis.

#### Feature selection

The feature selection step aims to reduce biases and inaccuracies in the features and also to minimize the feature space. In this step, we first removed the features with a large number of missing values. These features are difficult to impute and increase the sparsity of the modeling. Most of these removed features are missing completely at random (MCAR) or the underlying reason for missing values is difficult to deduce (i.e., MNAR) and have over 90% of values missing without any distinguishable patterns. Figure 2 illustrates the features with high and low missing values. In total, 55 continuous features and 18 discrete features were removed in this step. The features were excluded due to being empty, having low variance, or having missing value of 90% or more.

**Fig 2.**
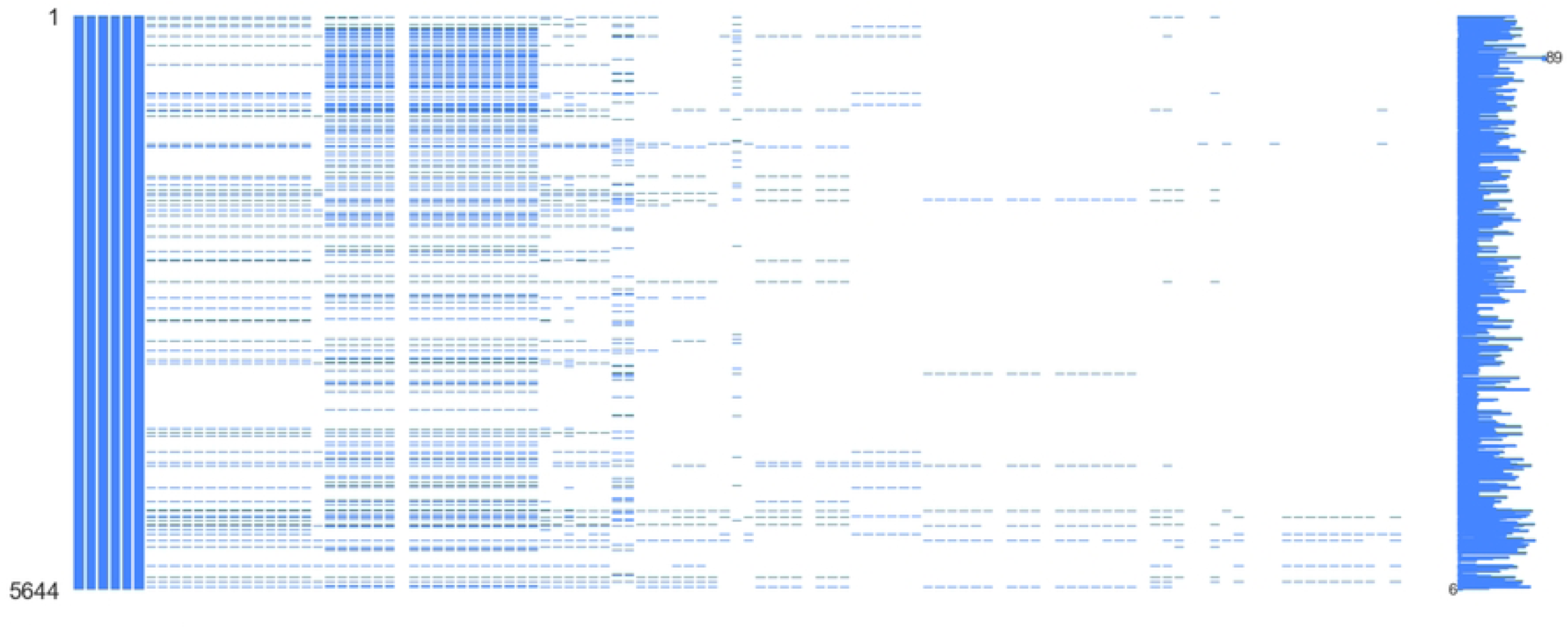
Missing features completely at random (MCAR). Blue color and white color depict features with complete and missing values, respectively.

Second, we use hierarchical clustering to identify test procedures that are closely related, therefore can be used to reduce the feature space. The clustering produces a clinically meaningful division of the data according to their similarities and patterns without requiring specification of an expected number of subgroups, as common clustering methods do [17]. The result is a division of features into three classes, which is used in the third phase as a basis for combining features. The categorization was verified according to clinical criteria to identify test families that are closely related. The resulting dendrogram is shown in Figure 3 and the clusters are as follows: (i) 14 blood count features that have been shown to be capable of detecting the presence of the SARS-CoV-2 virus at the second week of symptoms onset [18] and rapid tests for influenza A and B; (ii) 17 virus pathogen family variables; (iii) patient information including patient ID, the age quantile, SARS-CoV-2 exam result, and admission decision.

**Fig 3.**
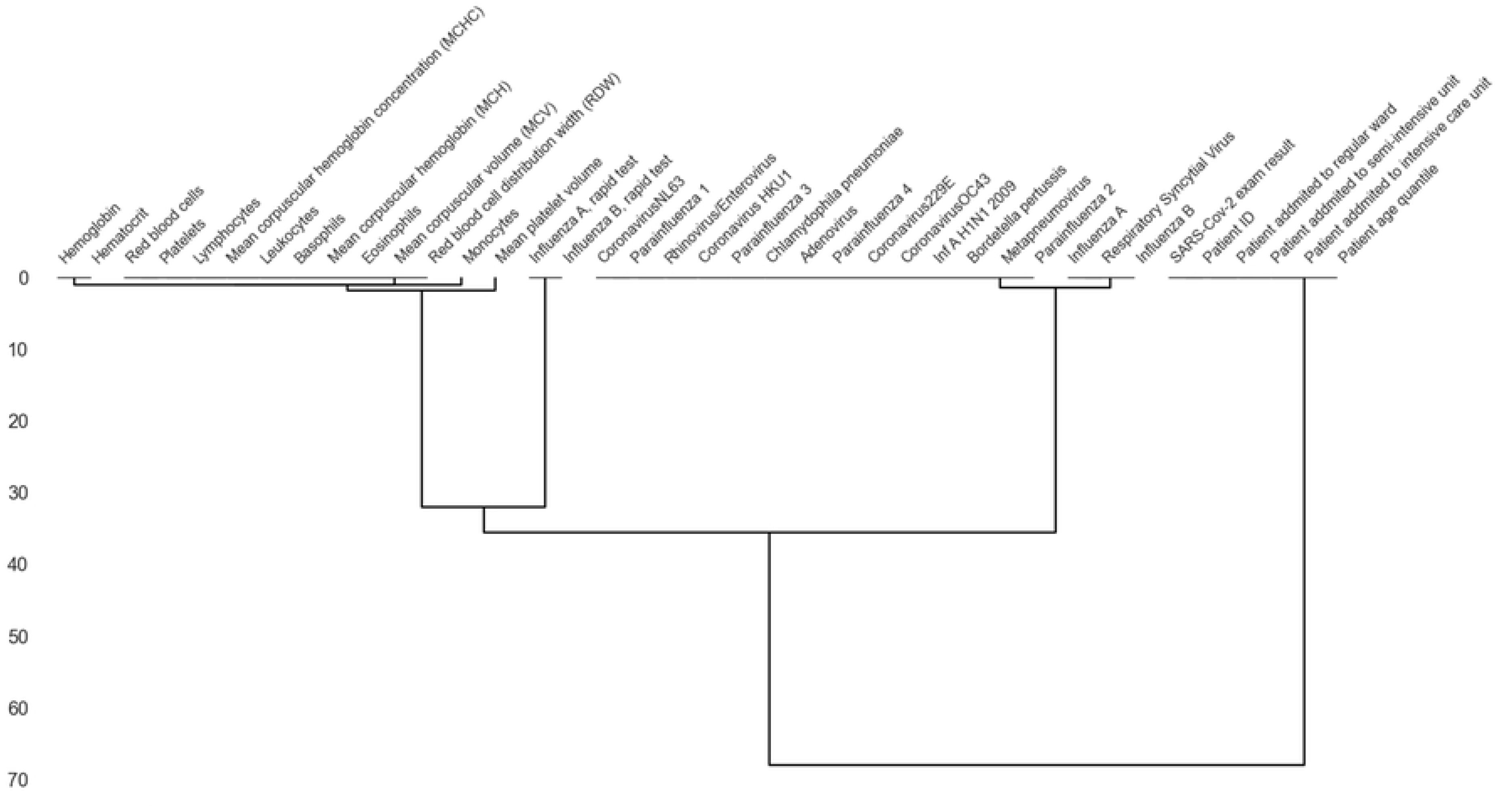
Dendrogram representing the feature clusters.

Third, we use the categorization resulting from the clustering in the second phase to merge features that are closely related. For this purpose, we create one variable for each virus family, reducing the feature space in the second cluster to six virus families. The general assumption is that the absence of pathogens has a likelihood of the patient not being infected. We encode each family with a binary variable indicating presence (1) or absence (0) of pathogens depending on whether any or none of the tests for a virus belonging to the family is positive. This produces the following six features:

1. *Adenoviridae:* Adenovirus
2. *Coronaviridae:* Coronavirusnl63, Coronavirus hku1, Coronavirus229e and Coronavirusoc43
3. *Orthomyxoviridae:* Influenza a, Influenza b and Inf a h1n1 2009
4. *Paramyxoviridae:* Parainfluenza 1, Parainfluenza 2, Parainfluenza 3 and Parainfluenza 4
5. *Picornaviridae:* Rhinovirus/Enterovirus
6. *Pneumoviridae:* Chlamydophila pneumoniae, Metapneumovirus and Respiratory syncytial virus

Additionally, we remove 2 rapid test features in cluster 1 from the feature space as they are already included in the Orthomyxoviridae virus family in cluster 2.

Finally, we analyzed features for redundancy which resulted in one modification to the feature space. Specifically, the feature hematocrit has a high correlation (96%) with hemoglobin and thus using only one of these features suffices. As the hemoglobin level is more common in the data, we remove the feature hematocrit from the feature space.

#### Labels

We used two labels for prediction. The first label is *COVID-19 diagnosis*, which is encoded as a binary variable where 1 shows having COVID-19 disease and 0 shows not having the disease. The second label is *potential patient deterioration* which its value obtained from the *admission to semi-intensive care units* and *admission to intensive care units* features. The label value is 0 if both *admission to semi-intensive care units* and *admission to intensive care units* are 0, otherwise the label value is 1. Note that this implies that we do not model the extent of deterioration, only whether the patient’s condition deteriorates or not. This means that patients admitted to regular ward (but not semi-intensive or intensive care) are not considered to have a deteriorated condition. This was done to improve the potential of AI/ML algorithms to learn robust dependencies from the data rather than fit on noise. Indeed, the number of patients admitted to semi-intensive or intensive care in the data is relatively small for modeling purposes.

### Missing data handling

The optimal way to handle missing data depends on the mechanism that results in the missing values, with the two main approaches being excluding the records with missing entries and replacing the missing entries using imputation [19]. The decision on whether to impute or not depends on the rate of missing values and the underlying mechanism that causes the missing entries. Generally, missing data can be a result of one of three processes: missing completely at random (MCAR), missing not at random (MNAR), or missing at random (MAR) [20]. MCAR is a result of random errors, such as equipment failures, whereas MNAR occurs when the mechanism causing the missing entries has no discernible patterns. MAR is the most common type of missing data and is defined as missing data that is dependent on observed values. Most of the missing data in our case results from one or more laboratory tests being omitted at the hospital depending on the medical analysis, and thus most of the missing entries in the data considered in this paper are MAR in nature. MCAR data can be deleted without risking adding bias to the results whereas imputation is recommended for missing values of MAR or MNAR types [21].

We separately assess how the profile of missing features affects the performance of imputation techniques and the performance of the AI/ML techniques used for predicting COVID-19 status and outcomes. The profile of missing features is characterized by the composition of the features that contain missing values (categorical, numerical, or mixed), the rate of missing values, and the pattern of the missing features. The pattern refers to the likelihood of data being missed together (togetherness) or independently (randomness). This pattern is motivated by the observation that certain combinations of laboratory tests either miss consistently or are present together. For example, features related to blood characteristics require a sample of blood from the patient and if this sample is not taken then all related features are necessarily missing. We verified the existence of this pattern by performing a pairwise correlation analysis of the presence and absence of features. All feature pairs with perfect correlation were merged into the togetherness pattern whereas others were encoded as random.

We assess four approaches for handling missing values: (i) excluding the missing data, i.e., no imputation (baseline); (ii) single imputation using a random draw from the observed values; (iii) multiple imputation using KNN; and (iv) multiple imputation using MICE (multiple imputation by chained equations). KNN and MICE are commonly used methods for imputing data [22] and they have been shown to perform well consistently, particularly for larger datasets [23].

The imputation techniques are evaluated using simulated datasets that are realistic but that reflect different profiles of missing data. As a baseline, we consider the complete data, and hence the evaluation is limited to the 362 samples that contain no missing values. We create 86 copies of these data by considering different levels of missing rate (5% – 90%, 1% increment) of three feature types (categorical, numerical, categorical+numerical) following the pipeline depicted in Figure 4. Missing rates of 5% and 90% are selected as a lower bound and an upper bound of the missing rate as clinical trial datasets having missing data less than 5% are classified as missing not at random (MNAR) whereas greater than 90% are considered as missing completely at random (MCAR), and cannot be meaningfully imputed [21]. Data is randomly dropped following either the togetherness pattern or the randomness pattern. In total, 516 datasets (3 feature types x 86 missing rates x 2 missingness patterns) were created in total, each represents a unique missingness profile. These datasets are referred to as simulated datasets. The three imputation techniques (single, KNN, MICE) were run on each simulated dataset 500 times and the mean absolute error (MAE, computed using Equation 1 of the values was computed and compared to the baseline, i.e., the complete data.

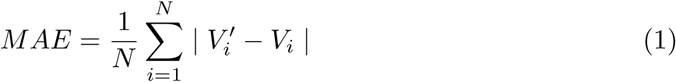

where N is the total samples, *V_i_* represents actual values and *V ^′^* denotes the values predicted by imputation.

**Fig 4.**
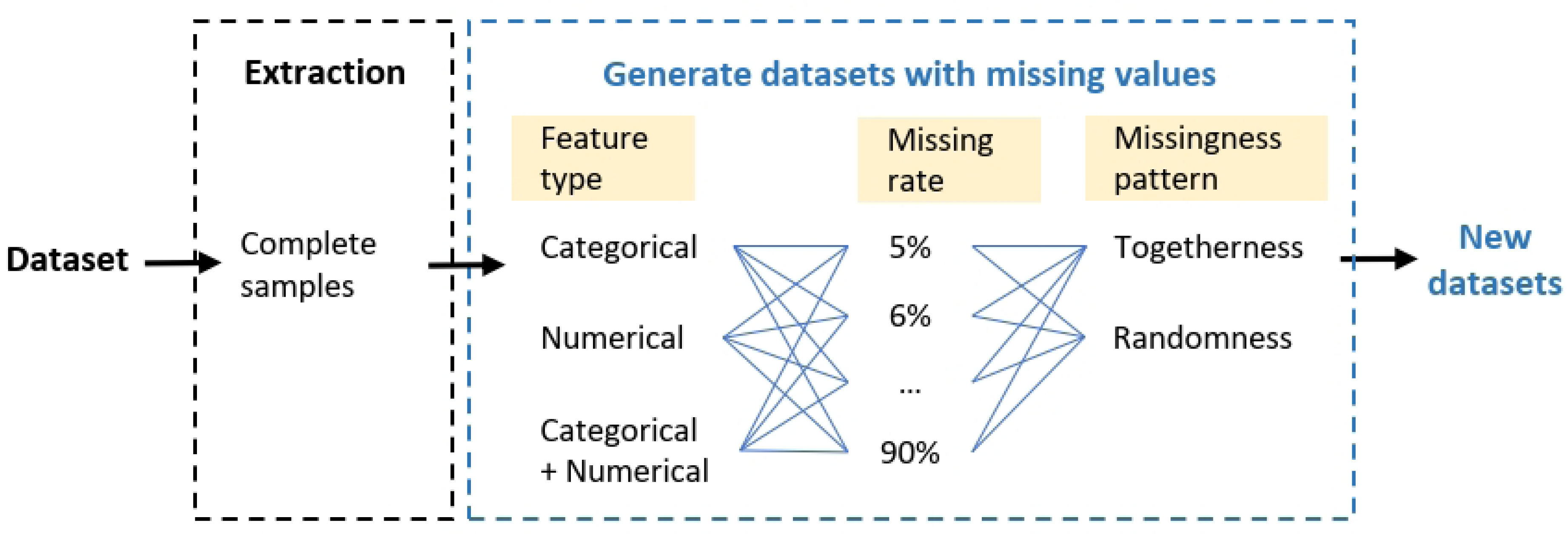
Pipeline generating datasets with different missingness profiles

### Infection and patient deterioration prediction

Classifiers were trained and tested for detecting COVID-19 infections and predicting patient deterioration from laboratory tests. As described in the previous section, the targets were encoded as positive (1) or negative (0) depending on the value of the RT-PCR test (Task 1) or whether the patient was admitted to semi-intensive ward or intensive care (Task 2). Both tasks were carried out on two datasets, one where all entries with missing values were excluded (i.e., complete) and one where entries with missing values were included; see Table 1. The latter was imputed using MICE as that yielded the lowest error in our analysis; see Section Imputation performance. Four different classification algorithms were considered: logistic regression (LR), random forest (RF), support vector machine (SVM), and k-nearest neighbors (KNN) models. These four learning models were selected for their performance on tabular data [24] and offering a degree of interpretability with feature ascription methods [25]. Deep learning was omitted due to the number of entries being insufficient for reliably validating overfitting.

**Table 1.**
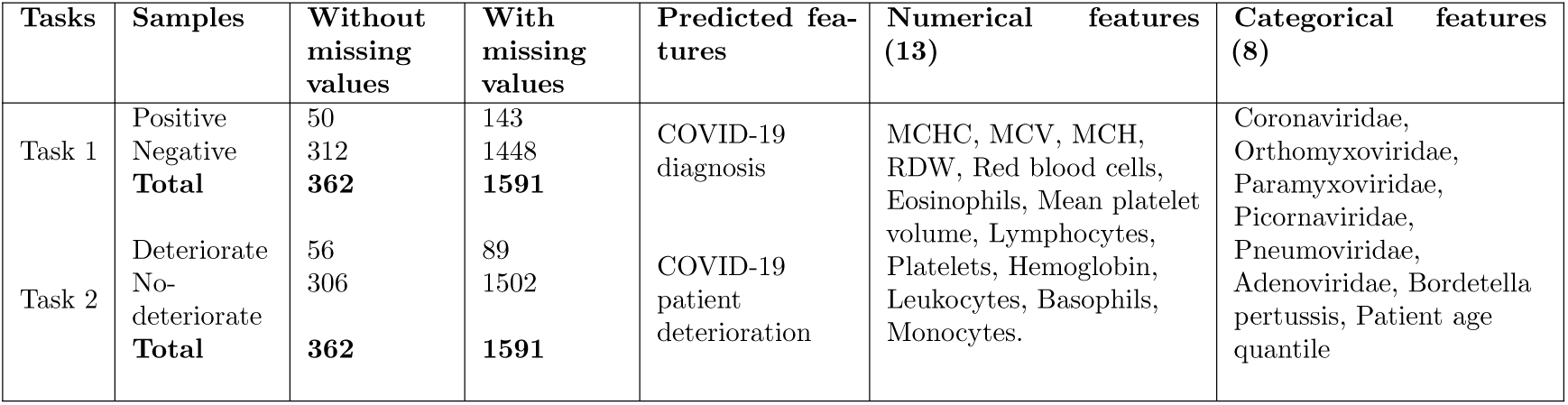
Dataset construction.

#### Evaluation metrics

The performance of the learning models is compared using accuracy, balanced accuracy, sensitivity, specificity, and the area under the curve (AUC). Sensitivity and specificity represent the percentage of individuals being correctly identified as positive or negative to the condition of interest, i.e., COVID-19 infection or patient deterioration, respectively. Sensitivity determines whether the test is satisfactory compared to a reference standard whereas specificity is important in establishing a patient tested positive on the screening test do or do not actually possess the condition [26]. Since the proposed imputation and AI/ML pipeline emphasizes both aspects, we calculated the average of sensitivity and specificity scores, i.e., balanced accuracy [13], and included it as a performance metric. Lastly, AUC depicts the degree of separability between classified classes, i.e., patients with and without COVID-19, or patients being admitted to semi-/intensive care units or not.

The training and test data were obtained using a stratified 5-fold cross-validation technique to balance the bias and variance of the small dataset during training. For each model, the best hyperparameter was searched and the best performing mode was saved for each case. To address the imbalanced class problem, the synthetic minority oversampling technique (SMOTE) was used to increase the minority class samples in the training dataset so that the model can generalize well on new unseen data. After applying SMOTE method, the dataset become more balanced to differentiate between the target classes.

To understand the impact of features on the predictive performance of the different models, we used Shapley Additive Explanations (SHAP) to estimate the average marginal contribution of each feature and use this to assess the importance of different features. The Shapley value originates from coalitional game theory and is a popular method for estimating the overall contribution of a feature for a given prediction [27]. The higher the absolute Shapley value of a feature, the more significant the contribution of a feature is on the predictions given by a model. The contributions of features can be visualized using a summary plot where the x-axis corresponds to the Shapley values and the y-axis to the different features.

### Implementation

All algorithms and experiments were implemented in Python (version 3.8). Numpy [28] was used for numerical data processing, Pandas [29] for data processing, Scikit-learn library [30] for both data processing and the classifier implementation, and Matplotlib [31] for data visualization.

## Results

In this section, we first present the performance of commonly used machine learning models on both complete and imputed datasets for infection prediction and patient deterioration prediction, evaluated in terms of accuracy, balanced accuracy, sensitivity, specificity, and AUC. Next, we investigate the important features for best model infection prediction and patient deterioration prediction using Shapley values. Finally, we compare our results with previous studies on COVID-19 detection.

### Prediction of COVID-19 infections and patient deterioration

The performances of predicting COVID-19 infections and patient deterioration are summarized in Table 2. The performance was assessed by comparing predictions from a complete dataset, where the records of patients lacking any laboratory test were removed from the dataset, to those from an imputed dataset. As shown in Table 2, all models have high accuracy and balanced accuracy, e.g., up to 97% and 90% in COVID-19 diagnosis, and up to 96% and 82% in predicting patient deterioration. The robust increment of these performance metrics as observed with imputed data demonstrates the effectiveness of proposed imputation and AI/ML pipeline.

**Table 2.** Results for predicting COVID-19 infection and for predicting patient deterioration. The best value for each score is denoted in bold.

| Dataset | Prediction | LR | RF | SVM | KNN | Average |
| --- | --- | --- | --- | --- | --- | --- |
| Complete | COVID-19 diagnosis |  |  |  |  |  |
|  | Accuracy | 0.91 | 0.91 | 0.90 | **0.93** | **0.91** |
|  | Balanced accuracy | **0.80** | 0.78 | 0.66 | 0.79 | **0.75** |
|  | Sensitivity | **0.64** | 0.59 | 0.32 | 0.59 | **0.54** |
|  | Specificity | 0.95 | 0.96 | **0.99** | **0.99** | **0.97** |
|  | AUC | 0.92 | 0.92 | 0.88 | **0.94** | **0.91** |
|  | Patient deterioration |  |  |  |  |  |
|  | Accuracy | 0.83 | 0.85 | **0.86** | 0.85 | **0.85** |
|  | Balanced accuracy | 0.74 | 0.78 | **0.79** | 0.77 | **0.77** |
|  | Sensitivity | 0.53 | **0.61** | **0.61** | 0.54 | **0.57** |
|  | Specificity | 0.96 | 0.96 | 0.97 | **0.99** | **0.97** |
|  | AUC | 0.69 | 0.74 | **0.84** | 0.78 | **0.76** |
| MICE Imputation | COVID-19 diagnosis |  |  |  |  |  |
|  | Accuracy | 0.96 | **0.97** | 0.96 | **0.97** | **0.96** |
|  | Balanced accuracy | 0.85 | **0.90** | 0.79 | 0.86 | **0.85** |
|  | Sensitivity | 0.71 | **0.81** | 0.58 | 0.72 | **0.70** |
|  | Specificity | 0.98 | 0.98 | **0.99** | **0.99** | **0.99** |
|  | AUC | 0.96 | 0.98 | 0.97 | **0.99** | 0.97 |
|  | Patient deterioration |  |  |  |  |  |
|  | Accuracy | 0.95 | 0.95 | **0.96** | 0.95 | **0.95** |
|  | Balanced accuracy | 0.81 | 0.80 | **0.82** | 0.76 | **0.80** |
|  | Sensitivity | 0.62 | 0.61 | **0.65** | 0.53 | **0.60** |
|  | Specificity | 0.99 | 0.99 | 0.99 | 0.99 | **0.99** |
|  | AUC | 0.79 | 0.84 | **0.94** | 0.90 | **0.87** |

#### Infection detection

The overall probability of correctly predicting COVID-19 infections is 70% and 99% for those without infections. All models have similar predictive performance, with an accuracy of at least 0.96, balanced accuracy of at least 0.79, and the minimum AUC score is 0.96. This means that those testing positive are unlikely to require further RT-PCR testing and thus our procedure can be used as a simple way to identify early infections prior to the onset of symptoms. In terms of algorithms, the RF model has the best performance, followed by KNN, LR, and SVM. Using RF on imputed data (sensitivity=0.81) shows an increase of 22% in correctly detecting COVID-19 infection cases compared to using it on the complete dataset. With SVM, this increment is 26% in correctly detecting COVID-19 infection cases from imputed data compared to complete dataset.

#### Patient deterioration prediction

As shown in Table 2, SVM is the best performing model for predicting patient deterioration, followed by LR, RF, and KNN model. The sensitivity score increases by 4% with SVM (sensitivity=0.65) and by 9% with LR (sensitivity=0.62) when the prediction is performed on the imputed dataset instead of the complete dataset Failing to detect the need for semi-/intensive care units for 9% of patients can rapidly burden the healthcare system and, in the worst case, lead to loss of life. While the sensitivity of the KNN and RF models remains practically unchanged, their AUC scores increase when data is imputed. This indicates a better ability to distinguish the patients that are likely to deteriorate and thus making imputation the preferred option. The specificity of all models is very high (minimum 0.96) regardless of whether the data is imputed or not. In other words, any positive cases for deterioration should be admitted to semi-/intensive care units as the likelihood of false positives is minimal. This highlights the potential of using our approach for triaging COVID-19 patients based on simpler laboratory tests.

### Feature importance

The feature importance in predicting COVID-19 diagnosis for the complete and the imputed datasets for the best performing models, i.e, RF using Shapley values is shown in Figure 5a and 5b, respectively. Figure 5c and 5d show the importance of features in predicting patient deterioration for the complete and the imputed datasets using the best performing model, i.e, SVM. These SHAP summary plots in Figure 5 consist of a horizontal bar chart where each bar in the plot corresponds to a specific feature. The length of the bar indicates the magnitude of the SHAP value, i.e., longer bars indicate features with a more significant impact on the prediction whereas shorter bars correspond to features with a lesser effect. The color of the bars indicates the feature values. For both models, all of the features have a similar impact on the model, with blood variables (leukocytes, platelets, monocytes, eosinophils) being consistently important for detecting the presence of coronavirus in the patient’s body.

**Fig 5.**
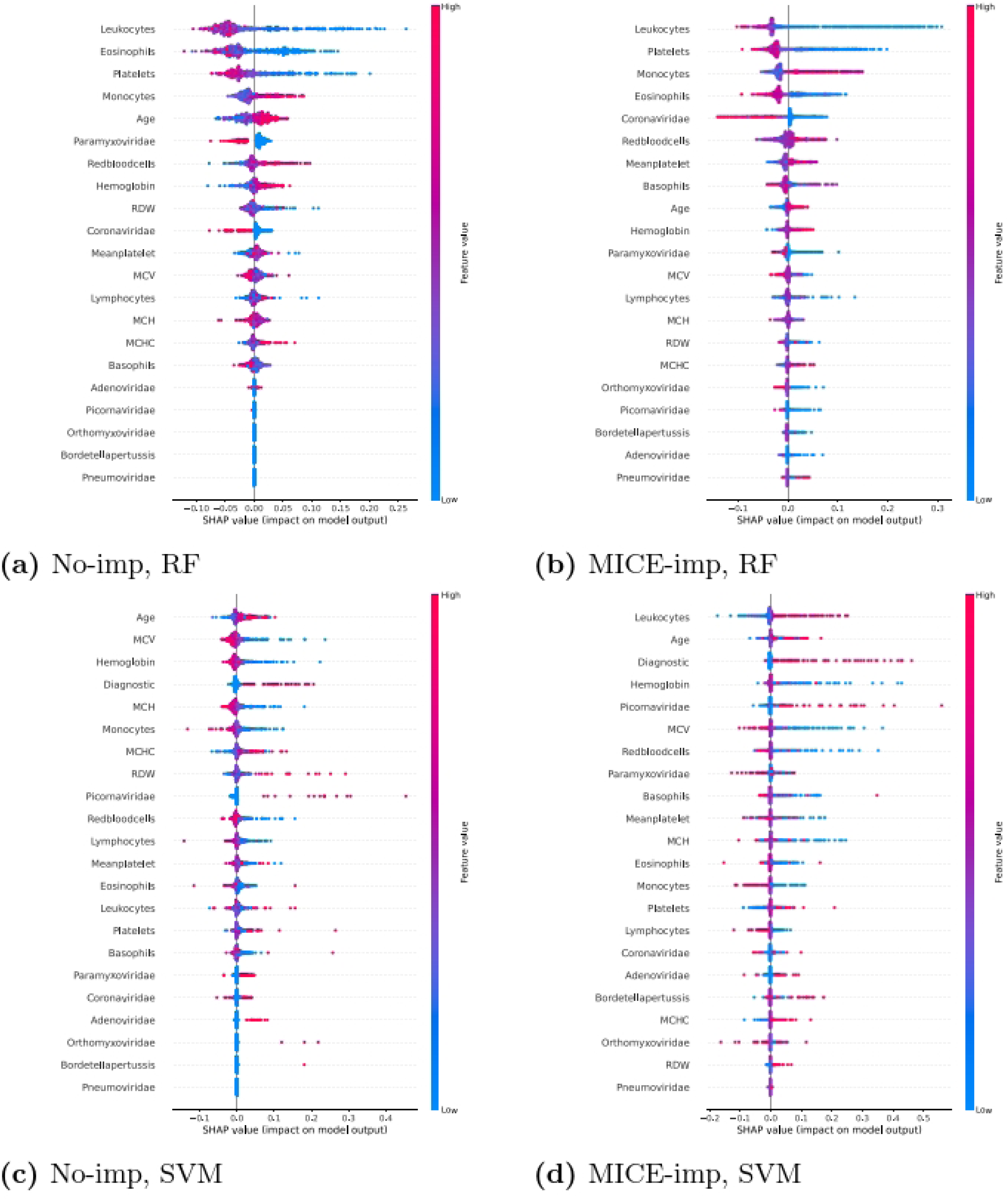
Impact of the features on predicting COVID-19 diagnosis (a, b) and patient deterioration (c, d).

### Imputation performance

In this section, we compare the performance of various imputation methods for different rates of missing data. Figure 6 depicts the mean absolute errors (MAE) of data imputation using simple, multiple imputation KNN, and multiple imputation MICE at 86 levels of missing rates (from 5% to 90% at 1% increments) of different feature types and patterns of missing data; following the simulated data generation pipeline detailed in Section Missing data handling. As shown in this figure, imputation using MICE yields lowest errors compared to KNN and simple imputation techniques regardless of the pattern or rate of missing data. Simple imputation is best suited for missing categorical data or data having high percentage of missing numerical data that are correlated. KNN imputation works better than simple imputation when the missing numerical data is uncorrelated, i.e., the values are randomly missing across multiple rows. When the missing data pattern is random, multiple imputation using MICE yields the lowest error. This holds up until more than 30% of the data has missing values, regardless of whether the data is numerical, categorical or a mixture of both. Most imputation techniques target data that is missing at random (MAR), and both single and multiple imputation also fare best in these cases with our data. KNN imputation seems best suited for data that is missing completely at random (MCAR). Overall, the results indicate that imputation is important for increasing data coverage, but that it is also important to assess the patterns of missing data and achieving the best possible result may require combining different imputation strategies.

**Fig 6.**
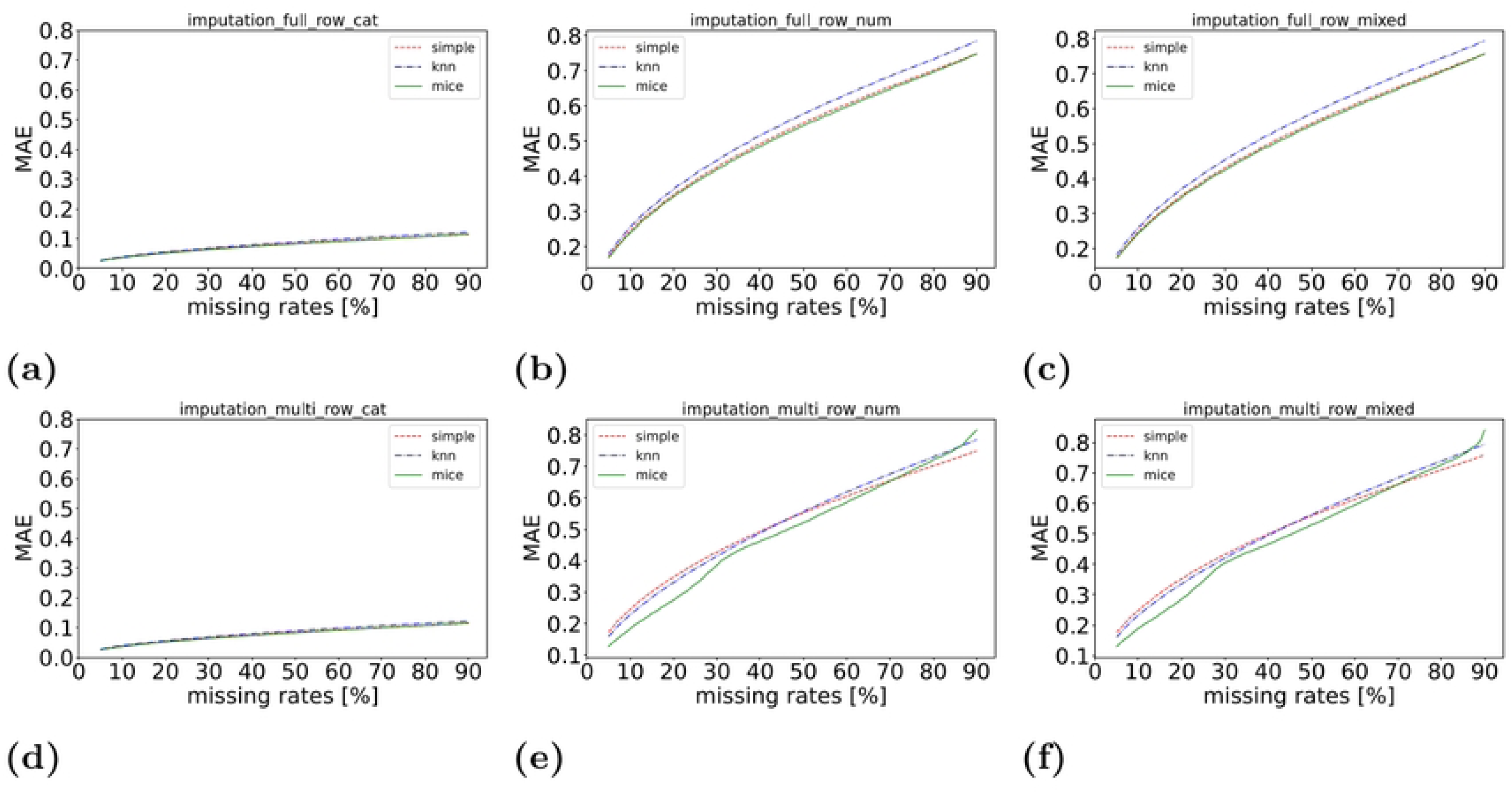
Mean absolute errors of data imputation in relation with missing rates (5%–90%) and missingness patterns (togetherness: a–c, randomness: d–f)

### Comparison with the previous work

Table 3 shows the performance of ML methods used in this study compared with the state-of-the-art methods for COVID-19 prediction. As indicated in the table, our models show higher specificity but lower sensitivity compared to prior research on using AI/ML for predicting COVID-19 infections or outcomes.

**Table 3.**
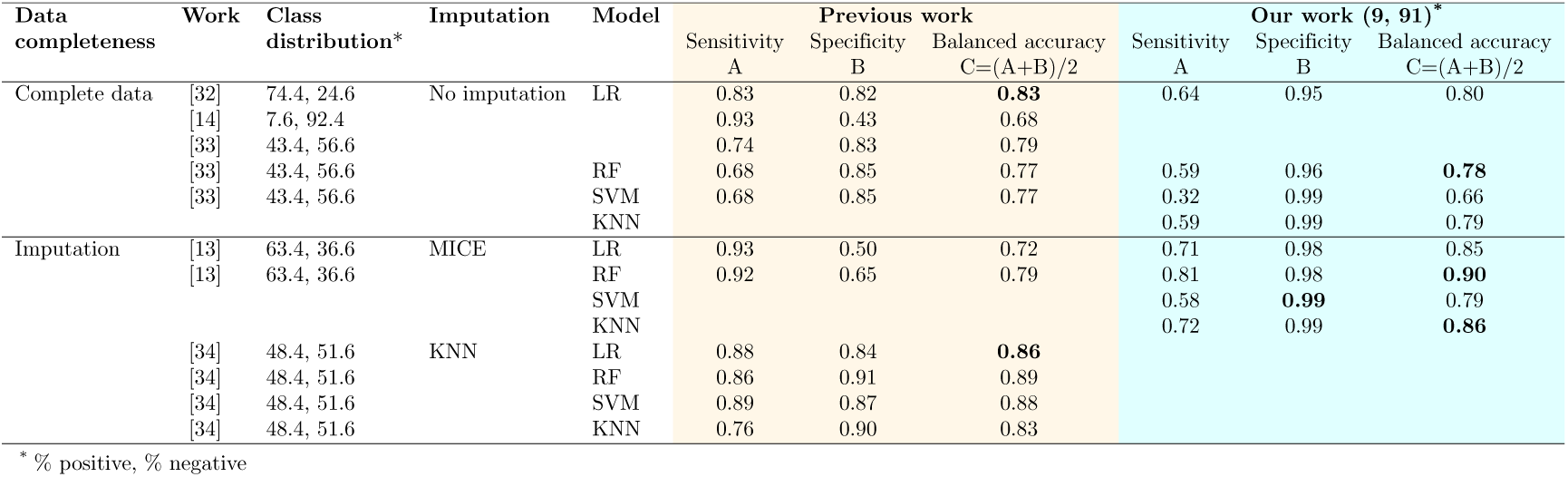
Performance comparison with prior works in COVID-19 diagnosis.

## Discussion

### Imputation and model performance

Handling missing data in clinical datasets is a significant challenge, as excluding records with missing values can introduce biases and lead to models with poor generalizability. Our study systematically investigates the effects of various imputation methods on AI/ML model performance, demonstrating that the choice of imputation strategy significantly influences the performance of ML models for detecting COVID-19 infection cases or patient deterioration. Specifically, we found that a lower number of infection cases are detected when missing values are excluded, which could result in many patients requiring admission to semi-intensive wards or intensive care units being overlooked.

Our results indicate that the accuracy, balanced accuracy, sensitivity, specificity, and AUC of different models for predicting both COVID-19 infections and patient deterioration are distinctly higher when using imputed data compared to a complete dataset, where records of patients lacking laboratory tests were removed. This demonstrates the importance of selecting appropriate imputation methods to ensure reliable clinical decision-making.

For effective early screening and triaging of infectious diseases like COVID-19, it is crucial to analyze features carefully prior to imputation and to impute only those where benefits can be realized. Our proposed approach yields better detection outcomes, particularly when features with less than 5% or over 90% missing values are removed to avoid biasing the models. In predicting patient deterioration, we found that imputing missing values significantly enhances prediction performance, highlighting the critical role of imputation in clinical applications.

### Clinical implications

The clinical implications of our findings are significant for multiple stakeholders. The AI/ML models for detecting infections leverage established laboratory procedures instead of requiring specialized tests, such as RT-PCR screening, which are often expensive and resource-intensive. As a result, our methods can facilitate timely interventions. Potential shortages of RT-PCR tests can hinder effective contact tracing and delay treatments. Early population screening using laboratory tests enables medical authorities and policymakers to understand disease prevalence and severity, allowing for effective prioritization and management of public health interventions. Additionally, demand prediction helps identify the needs of different hospital wards, alerting authorities to potential shortages of semi-intensive or intensive care unit beds in advance.

### Feature importance and its variability

In our analysis of feature importance, we observed that certain features, such as blood variables (leukocytes, platelets, monocytes, eosinophils), consistently play a crucial role in detecting COVID-19 infections. However, the importance of some features varies significantly depending on whether the data is imputed. For example, high levels of Pneumoviridae or low levels of Bordetella pertussis indicate a likely infection when the data is imputed, but these features do not contribute to prediction accuracy when trained on the complete dataset. This variability highlights the importance of imputation in exploiting correlations among features that may otherwise remain unutilized.

Similarly, in predicting patient deterioration, the significance of features changes after imputation. For instance, high values of leukocytes and Picornaviridae indicate a higher likelihood of deterioration, which is less apparent in the complete dataset. This emphasizes that the choice of imputation method not only affects model performance but also influences the clinical insights that can be derived from the data.

### Limitations and future work

While our study provides valuable insights into the effects of imputation methods on AI/ML model performance, it is not without limitations. First, the presented imputation and AI/ML pipeline aims at providing valid analytic estimation in the presence of missing data while preserving important relationships among features for predictive modeling rather than estimating the precise value for each variable. More advanced models for estimating the values of missing records could potentially further improve the early screening and triaging performance of AI/ML models. Second, the results are necessarily biased by the data which is provided by a single hospital in a single country and thus it is not possible to assess the generality of the methods to other populations or the impact genetic factors have on the algorithms. Nevertheless, the data set offers a systematically collected real-world dataset, and our results are consistently in line with medical studies that have analyzed effects of specific features on COVID-19. Third, our experiments assume that absence of pathogens indicates that the patient is not infected whereas in a real-world case the patient may not have been ordered for medical exams, e.g., as they are not showing any symptoms. Fourth, the results for the complete data have been calculated over a small patient cohort for which all medical tests were available. There can be external reasons affecting the availability of tests for these patients which cannot be assessed from the current data, and thus the results for the complete data may be biased and result in the machine learning models overfitting. The missingness profiles of the features indicate no clear correlations between features of different clusters, and the use of imputation improves the performance of the ML algorithms, indicating that any potential correlations or biases do not carry over from the complete data to the imputed data. To further generalize this imputation and AI/ML pipeline, future work may validate our proposed approach to datasets collected from different patient populations, medical and economical resources.

## Guidelines

The results demonstrated that both missing data and the selection of imputation techniques for handling it affect the performance of AI/ML models trained on patient records. In the following, we provide a summary of the key guidelines on how to best handle missing data to improve the quality of the data and AI/ML model performance.

**Step 1. Pruning the features.**

- *Feature-demand filtering:* The first step is to understand the nature and reason for missing data and to classify features with missing values as missing completely at random (MCAR), missing not at random (MNAR), or missing at random (MAR). The features classified as MCAR can then be omitted without the risk of adding bias into the results [12]. For the features classified as MAR and MNAR, only those having missing rates between 5% and 90% should be retained as otherwise they cannot be meaningfully imputed [21].
- *Feature-relatedness filtering:* Dependencies between features can degrade imputation performance and hence it is important to identify features (i.e., test procedures) that are closely related and to merge them. This can be done using e.g., hierarchical clustering, for producing a clinically meaningful division of the data according to their similarities and patterns.
- *Feature-redundance filtering:* Features that are highly correlated among one another contain significant redundancy and can again degrade imputation performance. As a result, only one of the redundant features should be selected with the one to retain being chosen based on the profile of the feature. For example, in case of blood tests, the information of the red blood cells can be obtained from hematocrit and hemoglobin test results, so selecting the one with the better feature profile suffices.

**Step 2. Identifying the missingness profile.** Features should be associated with a missingness profile that comprises the composition of the features that contain missing values, the rate of missing values, and the pattern of the missing features. In the context of personal medical records, the missingness profile can be determined based on the medical practices and procedures that govern which medical tests and procedures a specific patient undergoes.

**Step 3. Identifying the best imputation technique(s).** Imputation generally improves performance, but the imputation technique should be chosen based on the missingness profile of the feature identified in Step 2 and the type of the feature. As demonstrated in our experiments, *simple imputation* is best suited for missing categorical data or data having a high percentage of missing numerical data that are correlated. *KNN imputation* works better than simple imputation when missing numerical data randomly occurs across multiple rows. For data with a random missingness pattern, *multiple imputation using MICE* yields the lowest error.

## Conclusion

We systematically analyzed the effects of various imputation methods on the performance of AI/ML techniques, emphasizing the critical role of missing data handling in deriving accurate clinical insights. Our research highlights how the choice of imputation strategy significantly influences the performance of machine learning models, particularly in the context of COVID-19 detection and prognosis. We developed a cost-effective pipeline designed to differentiate between COVID-19 positive and negative patients while also predicting potential patient deterioration. Our findings demonstrate that the performance of machine learning models is heavily impacted by how missing data is managed, with improvements of up to 26 percentage points achievable through appropriate imputation techniques.

Utilizing four common classification algorithms, we investigated four strategies for handling missing data on a dataset containing medical records related to COVID-19 diagnosis and laboratory tests. Our results show that imputation consistently enhances predictive performance for the majority of classifiers compared to those using complete data, with only a few showing equivalent results. The Random Forest model (balanced accuracy = 0.90, AUC = 0.98) and the Support Vector Machine (balanced accuracy = 0.82, AUC = 0.94) were the top-performing models when using imputed data. These findings underscore the importance of addressing missing data, revealing that optimal imputation can lead to substantial performance enhancements. Importantly, the clinical insights derived from the models are contingent upon the imputation strategy employed, highlighting that the handling of missing data is critical for ensuring accurate interpretations and decisions in clinical practice.

Beyond providing guidelines and best practices for managing missing data, our work illustrates that effective handling of missing values is essential for robust AI/ML applications in healthcare. This is particularly relevant for healthcare systems in developing and underdeveloped countries, where access to advanced diagnostic tools may be limited. By leveraging AI/ML predictive models, these systems can enhance early screening and prioritize patients requiring immediate attention, even in the presence of significant missing data. However, the development of these models necessitates a thorough understanding of the effects of missing data and the strategies used to handle it, which our findings address. Ultimately, our research contributes to the advancement of AI/ML applications in healthcare, ensuring that accurate clinical insights lead to improved patient outcomes and more efficient resource utilization.

## Data Availability

The data used in this study is open-source and publicly available (released by Hospital Israelita Albert Einstein in Sao Paulo, Brazil) and can be accessed using this link: https://www.kaggle.com/datasets/einsteindata4u/covid19 The dataset is publicly available, and the publisher anonymized the patients data, removing all sensitive information, by following international guidelines. Researchers can use the data without any restrictions.

